# Face mask use in the general population and optimal resource allocation during the COVID-19 pandemic

**DOI:** 10.1101/2020.04.04.20052696

**Authors:** Colin J. Worby, Hsiao-Han Chang

## Abstract

The ongoing novel coronavirus disease (COVID-19) pandemic has rapidly spread in early 2020, causing tens of thousands of deaths, over a million cases and widespread socioeconomic disruption. With no vaccine available and numerous national healthcare systems reaching or exceeding capacity, interventions to limit transmission are urgently needed. While there is broad agreement that travel restrictions and closure of non-essential businesses and schools are beneficial in limiting local and regional spread, recommendations around the use of face masks for the general population are less consistent internationally. In this study, we examined the role of face masks in mitigating the spread of COVID-19 in the general population, using epidemic models to estimate the total reduction of infections and deaths under various scenarios. In particular, we examined the optimal deployment of face masks when resources are limited, and explored a range of supply and demand dynamics. We found that face masks, even with a limited protective effect, can reduce infections and deaths, and can delay the peak time of the epidemic. We consistently found that a random distribution of masks in the population was a suboptimal strategy when resources were limited. Prioritizing coverage among the elderly was more beneficial, while allocating a proportion of available resources for diagnosed infected cases provided further mitigation under a range of scenarios. In summary, face mask use, particularly for a pathogen with relatively common asymptomatic carriage, can effectively provide some mitigation of transmission, while balancing provision between vulnerable healthy persons and symptomatic persons can optimize mitigation efforts when resources are limited.

## BACKGROUND

The rapid global spread of SARS-CoV-2 and the resulting coronavirus disease (COVID-19) pandemic has led to urgent efforts to contain and mitigate transmission, leading to significant and widespread socioeconomic disruption. As of April 3rd 2020, over one million cases have been reported worldwide, as well as over 60,000 deaths, with ongoing spread in most parts of the world [1]. While infection is frequently asymptomatic, or associated with only mild symptoms in many people [2,3], it can cause severe and life-threatening illness in the immunocompromised and the elderly, with a case fatality ratio of over 10% in the latter group [3–5]. The rapid spread of the virus has raised concerns that healthcare systems lack sufficient resources and will be unable to bear the burden of accommodating patients suffering from COVID-19, resulting in significantly increased morbidity and mortality. There is an urgent need to better understand the effectiveness of potential interventions to limit the spread of the disease, especially in the context of resource limitations.

In order to avoid overwhelming national healthcare resources and mitigate the burden of infection, many countries have imposed both international and domestic travel restrictions, closed schools and non-essential businesses, and strictly limited public gatherings [6]. Such measures are designed to minimize person-to-person exposures, reducing the effective reproduction number R_0_, and thus the growth rate of the epidemic. Furthermore, behavior such as social distancing, self-isolation while symptomatic, handwashing, disinfecting surfaces and hygienic etiquette around coughing and sneezing can further mitigate transmission on an individual level. Interventions such as these can offer protection (reduction in risk of infection) to susceptible individuals, and/or containment (reduction in risk of onward transmission) to infected individuals. While such measures are near universally encouraged by governments and public health departments, there is limited international consensus on the use of face masks among the general public. Face mask use is common in East and South East Asia, and is currently recommended by the government in China, Hong Kong and Taiwan for healthy persons in crowded public spaces, as well as for symptomatic persons in Japan and Singapore [7,8]. Face mask use was compulsory in public in Wuhan, China, during public lockdown at the height of the epidemic. In contrast, the WHO does not currently recommend mask use among the general population [9], and the US government did not [10] until April 3, 2020, at which point cloth face coverings were recommended by the CDC [11]. There is a growing recognition across Western countries that face mask use should be part of public health policy for mitigating the spread of Covid-19. Recently, the Czech Republic made it mandatory to cover the nose and mouth in public [12], while Austria required the use of face masks in supermarkets [13].

Face masks may offer some degree of protection and containment; however, studies of mask effectiveness are relatively limited. While no studies to date have examined the effectiveness against transmission of SARS-CoV-2, the mechanisms of transmission via droplet and direct contact are likely highly similar to better characterized viruses. A cluster randomized household study examined the effectiveness of surgical and P2 (N95) masks in preventing transmission of influenza-like illness, finding a significant reduction associated with compliant mask use, although most participants failed to adhere to mask use sufficiently to see a benefit [14]. Cowling et al. showed that mask use and handwashing implemented within 36 hours of a case diagnosis could significantly reduce secondary household influenza cases (odds ratio 0.33) [15]. Mathematical models have also suggested that the number of influenza A cases can be reduced significantly even if just a small proportion of the population wear masks [16]. A systematic review on physical interventions to limit transmission of respiratory viruses, including severe acute respiratory syndrome, concluded that face mask use was effective [17]. In particular, N95 masks were slightly, but not significantly, more effective in reducing transmission than surgical masks, but were less comfortable to wear and caused skin irritation, potentially leading to lower compliance. Generally, the theoretical protective effect of masks may be diminished by a number of factors. Compliance and effective use may be inadequate [14], masks may not be replaced frequently enough to prevent contamination [18], and finally, Covid-19 infection may even occur via alternative routes, such as ocular transmission [19].

Although personal protection is a leading motivator for mask wearing [20], it is generally thought that face masks are more effective in providing containment, limiting onward transmission from infectious carriers. Surgical and N95 masks limit and redirect the projection of airborne droplets [21], and surgical mask wearing is estimated to be associated with a reduction in overall viral aerosol shedding [22] and coronavirus [23]. While diseases with a large proportion of symptomatic cases may result in most carriers reducing personal contacts (by choice or incapacity), Covid-19 is thought to have a high proportion of mildly symptomatic or asymptomatic cases, and therefore more infectious persons unaware of their status may continue to expose others. As such, even if masks offer limited personal protection, a general recommendation to wear masks in public may be particularly beneficial by containing transmission from unknowingly infectious persons.

Some countries have seen an enormous demand for face masks from the public, with supplies being diminished and shortages reported [7], drawing criticism for limiting supplies in healthcare facilities. Even with lesser public demand, the United States reported mask shortages among healthcare workers [24]. Recognizing the need for masks, the Central Epidemic Command Center (CECC) in Taiwan made efforts to increase mask production from January and announced when, where, and how to wear masks [25]. In facing such resource shortages, it is vital that limited supplies are used effectively. Clearly, protection of staff at healthcare facilities is of critical importance, but allocating additional resources optimally among the general population can offer further benefits.

In this study, we investigated the role of face mask use and distribution among the general public during a coronavirus outbreak, in order to better understand (i) the overall reduction in cases and deaths associated with mask distribution and use, (ii) how best to optimize distribution in a resource limited setting, and (iii) the role of dynamic supply and demand during an ongoing outbreak. With no available vaccine and limited options for treatment of COVID-19, it is crucial to allocate resources optimally to minimize cases and deaths worldwide.

## METHODS

In order to explore both the population-level effects of distributing facemasks to different subpopulations, as well as capturing the supply and demand dynamics during an ongoing epidemic, we proposed two models. Firstly, the **resource allocation model** allows a limited number of masks to be distributed among the initial susceptible population, or allocated to symptomatic individuals while supplies are available. This allows us to compare distribution strategies in terms of final numbers of infections and deaths. Secondly, the **supply & demand model** captures dynamic mask availability, which varies in response to increased demand among the entire population as the number of reported cases increase, as well as mask production rates.

Both models share the same basic epidemic SEIRD model structure (Figure 1) and the assumption of a closed, randomly mixing population of size *N*. Upon infection, susceptible individuals (*S*) enter the exposed (*E*) compartment in which a person is non-infectious. Infections progress to pre-symptomatic (*I*_*P*_) in which a person is infectious, but exhibiting no symptoms, and then either mildly symptomatic/asymptomatic (*I*_*A*_) or symptomatic (*I*_*S*_), similar to the model structure described by Anderson et al. [6]. Infected persons in either category can then recover (*R*), or if symptomatic, may die (*D*). Each compartment is partitioned into those wearing masks, and those not. Masks reduce susceptibility to infection in healthy individuals; mask wearers’ susceptibility relative to non-wearers is denoted *r*_*s*_, where *r*_*s*_=0 represents a fully protective mask. Similarly, masks are assumed to decrease transmissibility in infectious persons; a mask wearer has a relative transmissibility of *r*_*t*_, with *r*_*t*_=0 representing a mask completely restricting onward transmission. For convenience, we also define the terms ‘protection’ and ‘containment’ as 1– *r*_*s*_ and 1– *r*_*t*_, respectively, and use ‘mask effectiveness’ to refer to these properties collectively.

**Figure 1.**
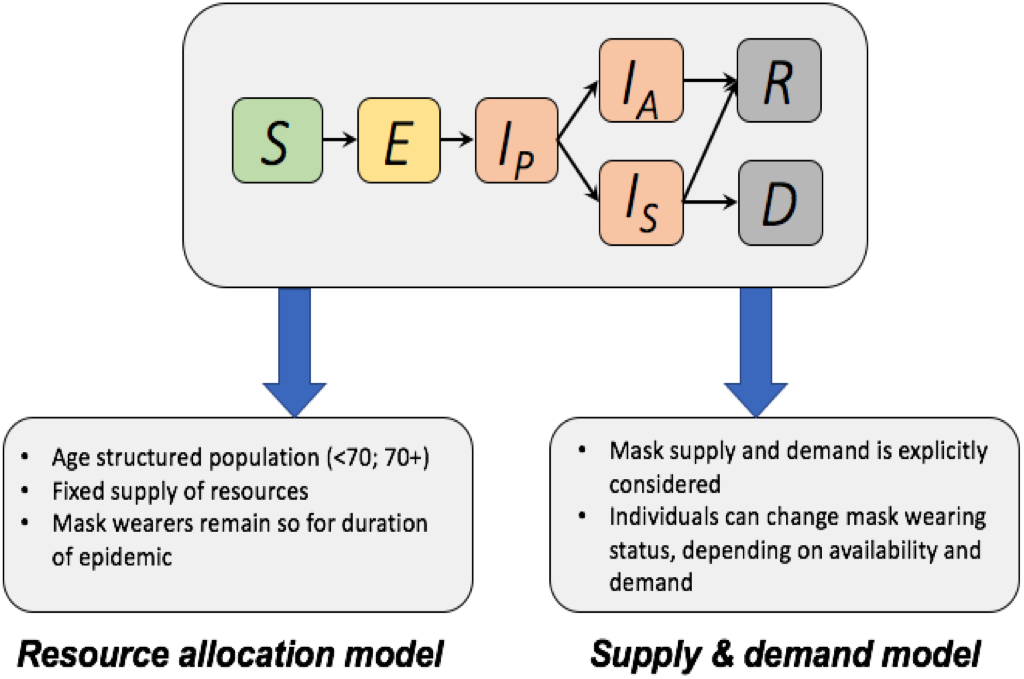
The compartmental structure common to both models used in this study is shown at the top. Susceptible hosts (*S*) become exposed (*E*) and progress to pre-symptomatic infectious (*I*_*P*_). Infected hosts can become either asymptomatic or mildly symptomatic (*I*_*A*_), or symptomatic (*I*_*S*_). Recovery (*R*) or death (*D*) follow. The resource allocation model and the supply & demand model then have unique additional features and dynamics. For a full description of each model and the specification of dynamics between compartments, see the Supplementary materials.

We obtained epidemiological parameter estimates from the available literature, and acknowledging the considerable uncertainty associated with many characteristics of disease spread, explored a range of values for certain key values. We assumed a basic reproduction number for symptomatic cases of R_0_=2.5 [6,26], and a 14 day period from symptoms to recovery [27]. Estimates for the proportion of asymptomatic cases vary considerably; including 18% based on data from the Diamond Princess cruise ship (with a high proportion of elderly people) [28], 25% according to the director of the US CDC [29], 30% based on Japanese evacuees from Wuhan [30], and even up to 78% based on limited reporting from China [31]. Given the importance of this parameter in determining allocation of resources, we allowed this parameter to vary over a plausible range. Mild/asymptomatic cases are assumed to have an infection rate 50% lower than fully symptomatic cases. While there is currently limited evidence on the progression of infection in different age groups, we assumed that the proportion of asymptomatic cases in the elderly (70+) was half the proportion in the younger population, and that the death rate among symptomatic elderly cases was 9.7%, versus 1.3% in younger cases [32]. All parameters used in the model are provided in supplementary table 1.

### Resource allocation model

In this model, the compartments described above are further partitioned by age group (young, <70, and elderly, 70+). We assume elderly persons are more likely to progress to symptomatic infection, and additionally are more likely to progress from symptomatic infection to death than young people. All symptomatic infections are assumed to be detected, while asymptomatic/mild infections are detected with a given probability. We assume a fixed supply of masks, sufficient to protect *M*_*0*_ persons in the population for the duration of the epidemic. Masks may be adopted by the healthy population at the start of the epidemic, while a certain proportion may be withheld for individuals with detected infection during the outbreak. We explore a variety of strategies to determine how masks may be optimally distributed such that total infections and deaths are minimized:

- **Strategy 1:** *M*_*0*_ of the susceptible population wear masks at the start of the epidemic.
- **Strategy 2:** *M*_*0*_ of the susceptible population wear masks at the start of the epidemic, with prioritized coverage of the elderly.
- **Strategy 3a:** 0.25*M*_*0*_ susceptible individuals wear masks at the start of the epidemic, prioritizing the elderly. Remaining masks are distributed to detected infectious individuals until supplies are diminished.
- **Strategy 3b:** 0.5*M*_*0*_ susceptible individuals wear masks at the start of the epidemic, prioritizing the elderly. Remaining masks are distributed to detected infectious individuals until supplies are diminished.
- **Strategy 3c:** 0.75*M*_*0*_ susceptible individuals wear masks at the start of the epidemic, prioritizing the elderly. Remaining masks are distributed to detected infectious individuals until supplies are diminished.
- **Strategy 4**: All available masks are distributed to detected infectious individuals.

For a range of mask effectiveness parameters, we identify the mask distribution strategy which minimizes both the number of infections and the number of deaths in the population. The model details are described in Supplementary Materials and baseline parameter values are listed in the Supplementary Table. While we do not explicitly model individual mask use and manufacture here, this can be thought of as continuous production to provide an equilibrium number of masks which may effectively be used by the fraction *M*_*0*_ of the population for the duration of the epidemic. A more explicit model of mask use is described in the following model.

### Supply & demand model

To understand the interplay between mask availability and disease dynamics, we modeled the supply and demand of face masks in a population, allowing masks to be produced at a given rate *B*, while demand may increase as the reported number of cases increases [7,33]. In this model, we allow for movement between mask-wearing and non-mask-wearing status, depending on availability and demand. Masks must be continually acquired to remain a mask wearer. We assumed that the mask is worn on average for *μ* days before requiring replacement, and the rate of non-wearers acquiring masks depends on both demand (*ω*_*A*_ for healthy and asymptomatic individuals, or *ω*_*S*_ for symptomatic individuals) and current supply (*M*/*N*, the proportion of mask in the overall population). We assume that demand for masks increases with the number of reported cases in the population up to a certain plateau, and as such modeled the relationship between *ω*_*A*_ and the number of symptomatic infections in the following sigmoidal function:

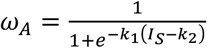

The range of *ω*_*A*_ is [0, 1], *k*_*1*_ represents the rate of demand increase, and *k*_*2*_ represents the timing of demand, defined as the number of reported cases when half the population seeks face masks (i.e. higher *k*_*2*_ means mask demand increases later in the outbreak). We allow *ω*_*S*_ to differ from *ω*_*A*_, in order to explore the effects of recommending face mask use to the general population (*ω*_*S*_=*ω*_*A*_), or specifically to symptomatic individuals (*ω*_*S*_>*ω*_*A*_). This parameterization allows us to explore different demand dynamics, for example, panic buying (high *k*_*1*_), delayed response to epidemic threat (high *k*_*2*_), limited interest in mask use (low *k*_*1*_, high *k*_*2*_) (Figure S3).

## RESULTS

### Resource allocation model

We simulated outbreaks under a variety of parameter values associated with mask effectiveness (protection and containment) and mask supply, and identified the resulting total numbers of infections and deaths under each strategy described in the methods. Figure 2 shows the impact of mask distribution under different strategies, for different levels of availability. Reduction in total deaths increased with mask effectiveness and availability. Even limited distribution of relatively ineffective masks could result in an appreciable reduction; 10% adoption in the population could result in 5% fewer deaths (Fig. 2, top right). While immediate provision to the healthy population provided maximal impact, delayed implementation of a general mask wearing policy could still provide reductions in total cases. The epidemic peak could be increasingly delayed with earlier adoption of mask use (Figure 3).

**Figure 2.**
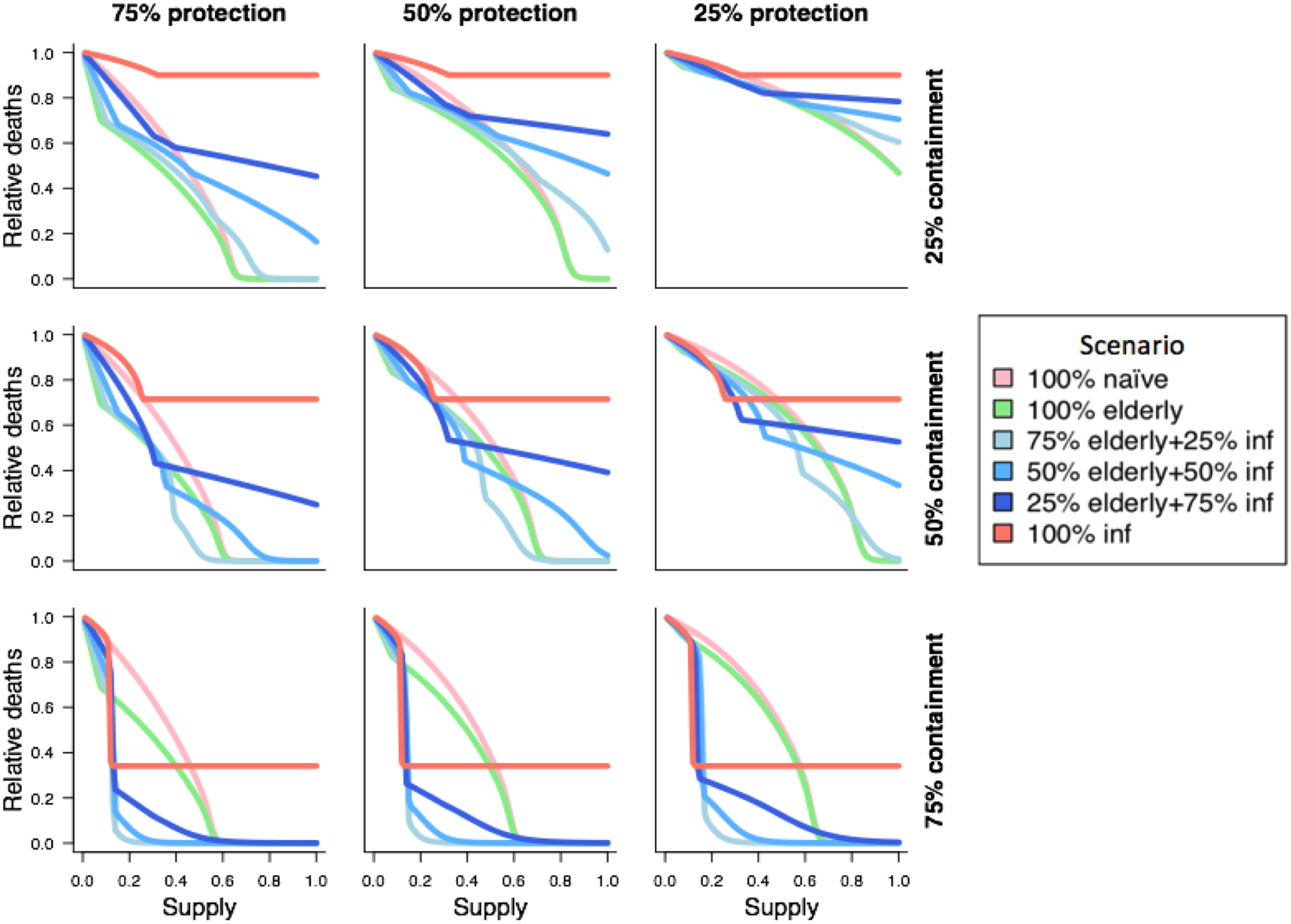
Reduction in total deaths under each of the described resource allocation strategy for a range of resource availability levels. Each panel represents intervention effectiveness in terms of relative susceptibility and transmissibility, with the bottom left panel denoting the most effective intervention (75% reduction in susceptibility and transmissibility) and the top right panel representing the least effective intervention (25% reduction in susceptibility and transmissibility). Resources are provided naïvely (pink), prioritized to the elderly (green), saved for detected cases (red), or balanced at different levels between healthy individuals, prioritizing the elderly, and detected cases (blue). 30% of cases are assumed to be undetected. See *Methods* for further details.

**Figure 3.**
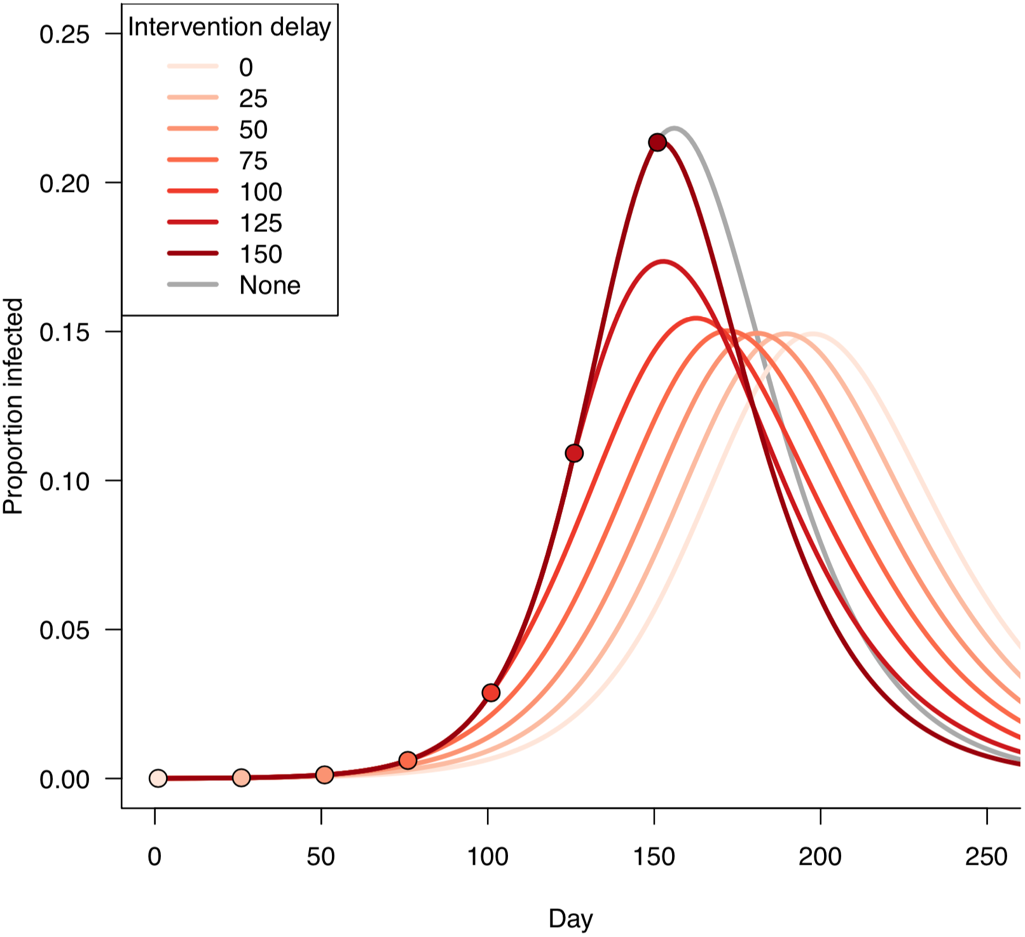
Rapid introduction of face masks to the general population can reduce cases and delay the epidemic peak. For a range of implementation dates, 25% of the general population adopt face masks conferring 25% protection and 50% containment.

Naïve distribution of masks among the general population (strategy 1) was usually suboptimal; indeed, for a mask providing better containment than protection (*r*_*t*_<*r*_*s*_), this is the least optimal of the strategies we tested unless resources were plentiful (Figure 2, bottom right panels). While prioritizing allocation to elderly persons (strategy 2) only slightly reduced the total number of infections beyond that achieved with naïve distribution (Figure S1), the number of deaths was generally much lower with this strategy. The benefit of prioritizing the elderly population was largest in scenarios with a limited supply of protective masks, diminishing gradually with masks offering more limited protection. With plentiful resources, the difference between prioritizing the elderly population and random distribution became limited.

Providing masks only to detected cases was an effective strategy in some limited resource scenarios when containment was high (Figure 2, red lines). However, since many infections are not detected, this strategy fails to provide any containment to the large, undetected infective population, and the benefits associated with increasing supply reach a maximum once there are sufficient resources for all detected cases. As such, this policy offers the least optimal distribution for a range of mask effectiveness parameters when resources are abundant (e.g. >50% of the population can be covered). As an intervention that offers no protection to susceptible individuals, the associated reduction in total infections only depends on mask containment. By providing a mask offering intermediate levels of containment (50%) to all detected infectious cases, the number of deaths can be reduced by up to 30%, reaching this level with resources to cover 25% of the population. Increasing the case detection rate can further increase the benefits of this strategy.

In many scenarios, achieving a balance between providing resources to infective persons and the elderly population offers the optimal outcome in terms of total infections and deaths (Figure 2, blue lines). These strategies (3a-c) offer containment focused on symptomatic cases, but also mitigating transmission from a proportion asymptomatic carriers. Additionally, some degree of protection is granted to susceptible individuals, with a focus on the more vulnerable elderly population. Figure 4 shows the optimal distribution strategy over the full range of mask effectiveness parameters with resources to cover 40% of the population, as well as the corresponding reduction in total infections and deaths, relative to the naïve strategy of random distribution. The optimal balance of mask distribution varied according to supply and mask effectiveness. Generally, while resources are plentiful, providing the majority of available supplies to the healthy population (prioritizing the elderly) at the start of the epidemic was optimal (e.g. strategy 3a) for reducing cases and deaths. Increasing the case detection rate among mild/asymptomatic cases will improve the effectiveness of these balanced strategies.

**Figure 4.**
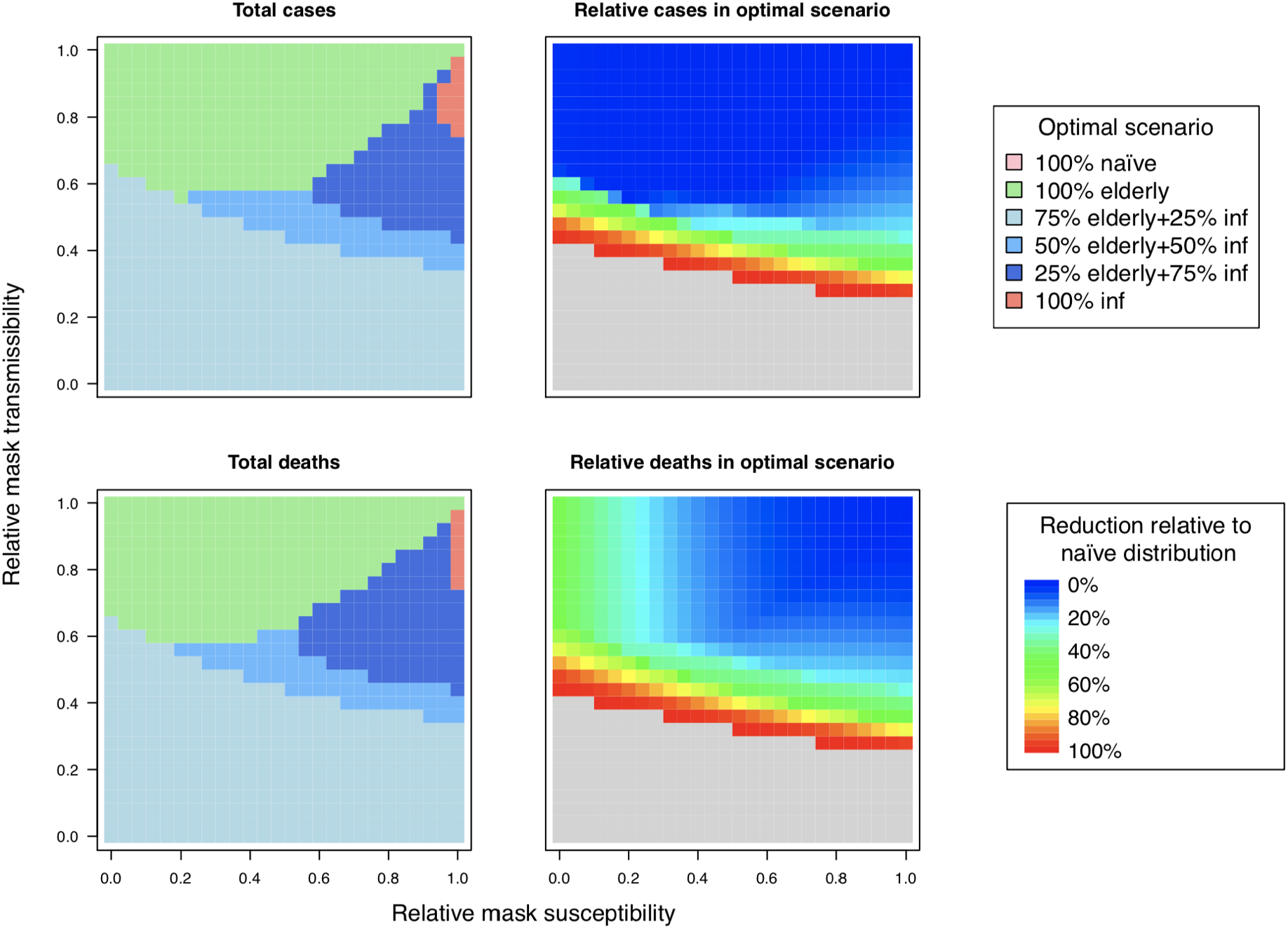
Optimal distribution of resources for different levels of intervention effectiveness. The strategy which minimized the number of infections (top left) and the number of deaths (bottom left) is indicated for each level of intervention protection and suppression. With a supply of masks for 40% of the population, resources are provided under each of the strategies described in *Methods*. The reduction in infections and deaths under the optimal strategy is shown in the right column, relative to the numbers under the naïve strategy. The area shaded in gray represents complete suppression of the outbreak under the optimal strategy. Here we assume the probability of asymptomatic carriage to be 0.3 among cases <70y.

Optimizing mask distribution offering limited protection and containment unsurprisingly had a minimal additional benefit beyond random distribution in the healthy population (Figure 4, right). However, we found that while total infections remained similar, optimizing distribution had the effect of delaying the peak week of the outbreak (e.g. Figure S2). In practice, this is highly desirable for providing additional time to scale up healthcare resources and infrastructure, and may result in an indirect reduction of morbidity and mortality associated with overstretched healthcare facilities.

A far smaller supply of highly effective masks were required to achieve similar reductions in total infections compared to a large supply of less effective masks. Masks retained for provision to infectious persons are more rapidly exhausted when they provide either limited protection or containment. Deaths could be reduced by 65% with 15% coverage of a highly effective mask (75% containment), compared to a reduction of 30% with 25% coverage of an intermediate mask, and a reduction of 10% with 30% coverage of a low effectiveness mask (25% containment). In the resource allocation model, new mask production and ongoing supply is not explicitly considered. In the following model, we investigate the role of these dynamics.

### Supply and demand model

We explored different models of mask availability and demand by varying the parameterization of the demand function described in the methods, as well as the rate of production of new masks. Unsurprisingly, regardless of demand dynamics, a higher production rate of masks increased availability and therefore coverage of the population. Figure S4 shows the reduction in total infections given different levels of protection and mask production, highlighting that a greater number of less effective masks were required to achieve the same impact as fewer, but more effective, masks. For a given level of mask production, a ‘panic buying’ scenario, in which maximum demand for masks was attained very early in the epidemic, generally had a detrimental impact on the resulting outbreak (Figure S5). Unless production is ramped up during the outbreak, an inability to build a stockpile of available resources prevents infected persons obtain masks readily during peak transmission (Figure 5, left). In contrast, a more gradual increase in demand, or equivalently, a managed distribution of resources such that a stockpile can be built up in the early stages of the epidemic, results in greater availability of masks during peak transmission, and fewer overall cases (Figure 5, right). Prioritizing provision to infectious cases (setting *ω*_*S*_>*ω*_*A*_) can reduce cases further (Figure 5, bottom), in line with our findings from the resource allocation model, under the assumption that masks offer a greater degree of containment than protection. This effect is smaller if the proportion of asymptomatic infections is high (Figure S6). While building up supplies in the early phase of the epidemic can be beneficial, high levels of production were required to avoid shortages during peak demand (Figure S7).

**Figure 5.**
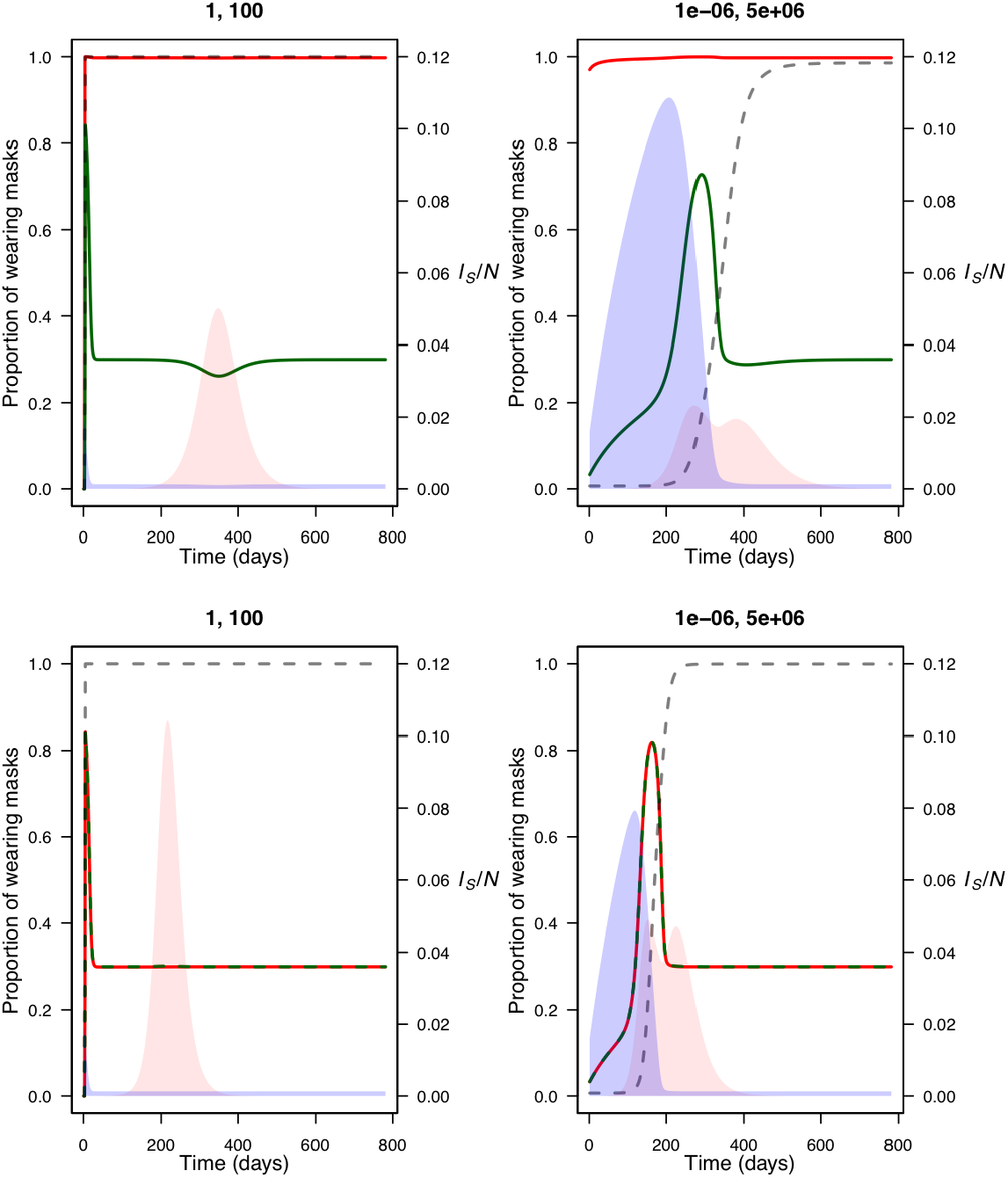
Managing resource demand in the early stages of the outbreak can limit the total number of infections. Epidemic curves (pink) under a ‘panic buying’ demand curve (left), and a more gradual managed demand curve (right) when prioritizing (top) and not prioritizing (bottom) masks for infectious cases. Demand is shown as a dashed grey line, while the relative available mask supply is shown in blue. The proportion of the susceptible and symptomatically infected population wearing masks are shown as green and red lines, respectively. (*k*_*1*_, *k*_*2*_) are shown on the top of each subfigure. The dynamics shown here are based on mask production rate, *B*/*N*, equal to 30%.

## DISCUSSION

During a pandemic such as COVID-19, mitigating the spread of infections is essential in the absence of a vaccine and limited critical care resources. In this study, we have shown that face mask use in the general population can have a beneficial impact in reducing the total number of cases and deaths, and that this impact naturally increases with mask effectiveness. The benefits of mask deployment are apparent even with low effectiveness and limited resources. In such cases, though mask deployment may not have a large impact on total cases and deaths, indirect benefits for outbreak management are achieved by delaying the epidemic peak. Importantly, however, the overall impact of mask deployment hinges on appropriate distribution strategies. We consistently observed that the random distribution of masks throughout the general population is a suboptimal strategy. In contrast, prioritizing the elderly population, and retaining a supply of masks for identified infectious cases generally leads to a larger reduction in total infections and deaths than a naïve allocation of resources. While there remains much uncertainty around the true effectiveness of face masks - especially when factoring in differences in mask types, levels of adherence and patterns of human behavior - there is evidence to suggest that masks can provide a measure of protection and containment for respiratory viruses, likely a greater degree of the latter than the former.

The more effective a mask is, the fewer masks are required to suppress an epidemic. Under a strategy in which masks are retained for infectious persons, this is particularly important. As a higher proportion of infectious persons - both symptomatic and asymptomatic (and possibly unaware of their status as a carrier) - are wearing masks offering a high level of containment, a smaller number of onward transmissions occur, requiring fewer masks to be provided for newly diagnosed individuals. While mask use can help to mitigate transmission, the supply & demand model suggests that panic buying at the very early phase of an epidemic can be detrimental, and that managing demand in the early stages of the outbreak could be beneficial. In Taiwan, the government implemented such a resource management strategy in early February 2020, limiting the number of masks each person can buy per week with their National Health Insurance cards [25]. As human behavior and compliance are a significant component of how effective mask use is, it is essential that public health recommendations concerning face masks in the general population occur in tandem with clear education on proper use and application, such that limited resources are used as effectively as possible.

During a pandemic such as COVID-19, optimizing the deployment of resources is essential. Our models concern the distribution of resources in the general population, under the assumption that healthcare workers and key personnel have adequate protection - clearly it is essential that this subpopulation should have prioritized access to protective equipment. However, if production can be increased such that face masks may be available to the general population, an optimized deployment of these resources can limit spread. While we considered the elderly population in our model, we did not explicitly consider other vulnerable subpopulations, such as immunocompromised persons or those with other respiratory conditions. Such persons with increased risk of suffering severe infections, and perhaps more importantly, those interacting with them, should belong to the prioritized subpopulation we considered in the resource allocation. In addition, we did not consider heterogeneity in population mixing. In reality, there are clusters of particularly vulnerable persons (e.g. hospitals, nursing homes, long term care facilities, prisons, homeless shelters) which pose an elevated risk; failing to protect such communities could lead to rapid and highly localized spread. It is likely that face mask use is also more beneficial in populations with higher contact rates. Future modeling work could consider meta-populations of different population densities to optimize resource deployment in urban vs. rural settings.

The use of face masks in mitigating the spread of a pandemic disease such as COVID-19 is one of many strategies that are being implemented simultaneously, including social distancing, travel restrictions and self-isolation. Even during lockdown measures in which people are only rarely leaving their homes, many still face high exposure settings (e.g. conducting essential work, trips to the supermarket) albeit less frequently. Face mask use could be a particularly important component of transmission mitigation once social distancing measures are relaxed, and potential exposures rapidly increase. Preparing an adequate supply of face masks for such a transitionary period could help to prevent a potentially costly second peak.

## Data Availability

N/A

## Funding

This study was supported by the Ministry of Science and Technology in Taiwan (MOST 109-2636-B-007-006). CW received support from the National Institute of Allergy and Infectious Diseases (grant number U19AI110818). The funders had no role in preparation of the manuscript.

## SUPPLEMENTARY MATERIALS

### Resource allocation model

Consider a population of size *N*, comprising *N*_*y*_ and *N*_*o*_ young (<70 years old) and elderly (70+ years old) respectively. At a given time, each individual has a status as a mask-wearer (*m*) or a non wearer (*n*). For each age group *x* (*x*=*y* for young, *x*=*o* for elderly), and status *s* (*s*=*m* for mask wearer, *s*=*n* for non wearer), let *S*^*xs*^, *E*^*xs*^, be the number of susceptible and exposed (but not infectious) individuals. Let 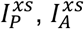 and 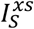 be the number of pre-symptomatic, asymptomatic/mildly symptomatic and fully symptomatic individuals respectively. Finally, let *R*^*x*^ and *D*^*x*^ be the number of recoveries and deaths. Fully symptomatic cases are associated with an infection rate of *β*_*S*_, while pre-symptomatic and mildly/asymptomatic cases have a lower infection rate *β*_*A*_; we assume *β*_*S*_ = 2*β*_*A*_. Progression from exposed to pre-symptomatic occurs at rate *α*_1_, and progression to either *I*_*S*_ or *I*_*A*_ occurs at rate *α*_2_. The probability of developing only mild symptoms or no symptoms among age group *x* is *P*_*ax*_. Removal occurs at rate *γ*_*S*_ and *γ*_*A*_ respectively. The proportion of fatalities among fully symptomatic cases in age group *x* is assumed to be *p*_*xD*_. Dynamics are governed by the following system of differential equations:

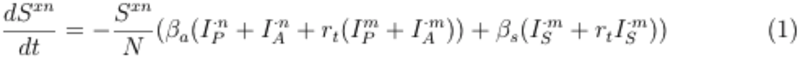

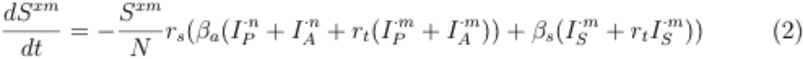

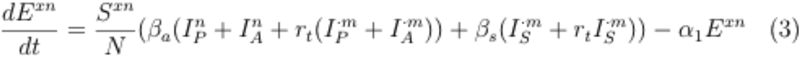

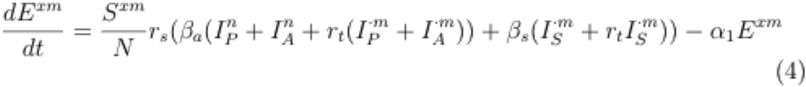

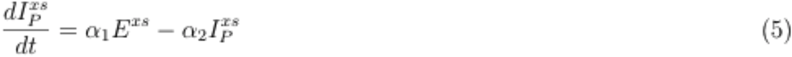

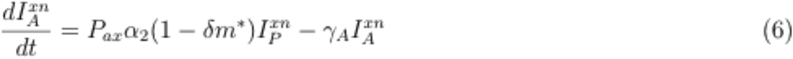

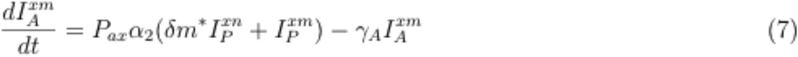

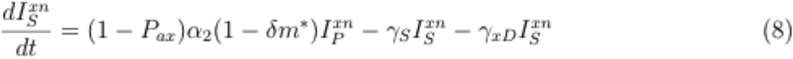

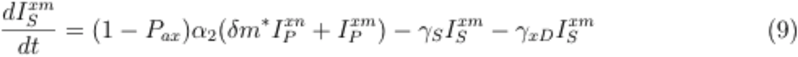

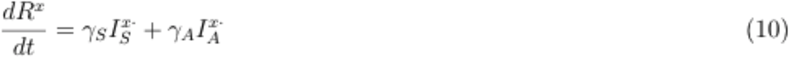

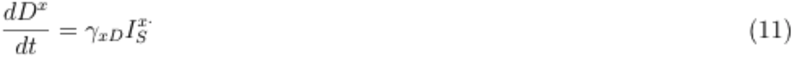

where *m\** is an indicator function equal to 1 when there is a remaining supply of masks *M*, and is the probability of detecting mild/asymptomatic infections. As masks are provided to newly diagnosed cases, supplies are depleted at the following rate:

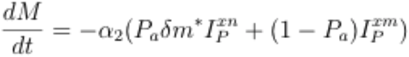

Monotonically declining until zero is reached, at which point, no further masks are provided for detected cases.

### Supply & demand model

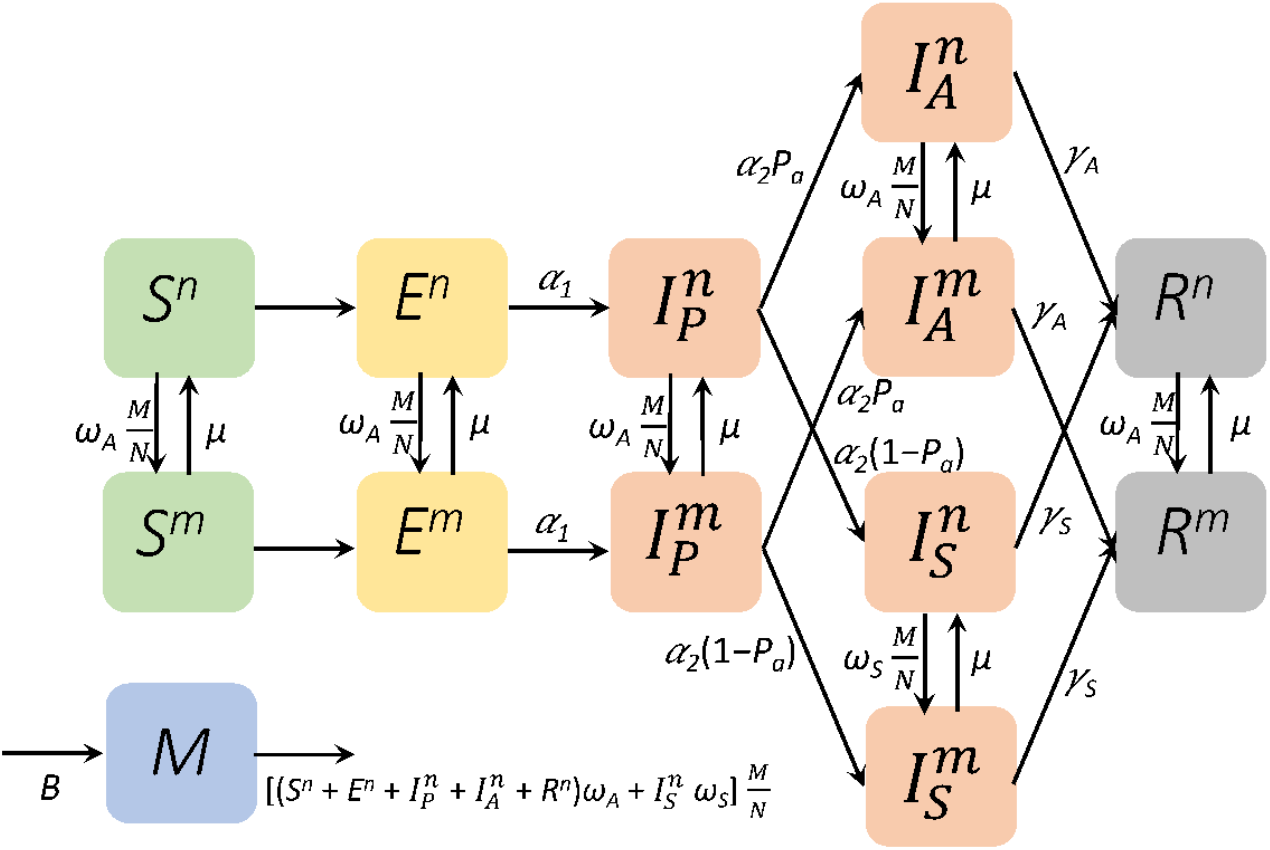

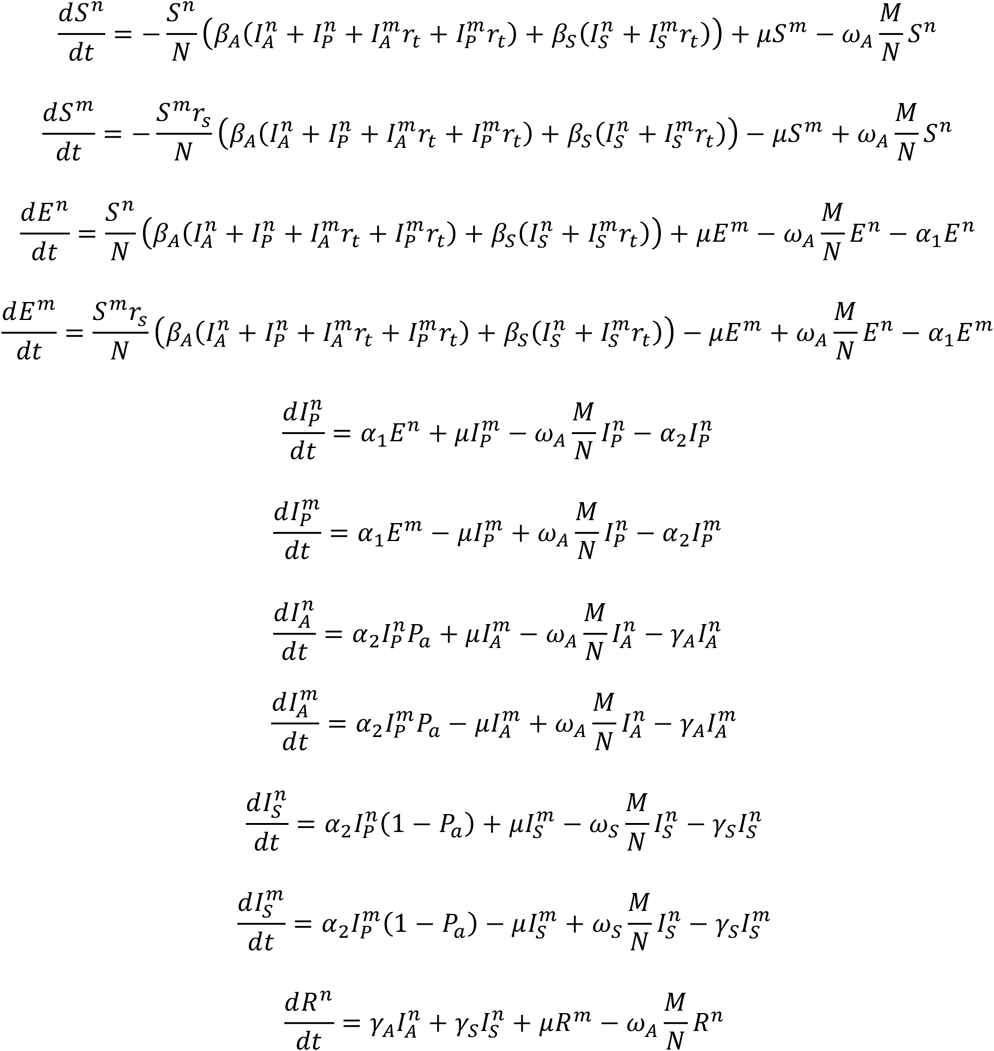

## SUPPLEMENTARY FIGURES

**Figure S1.**
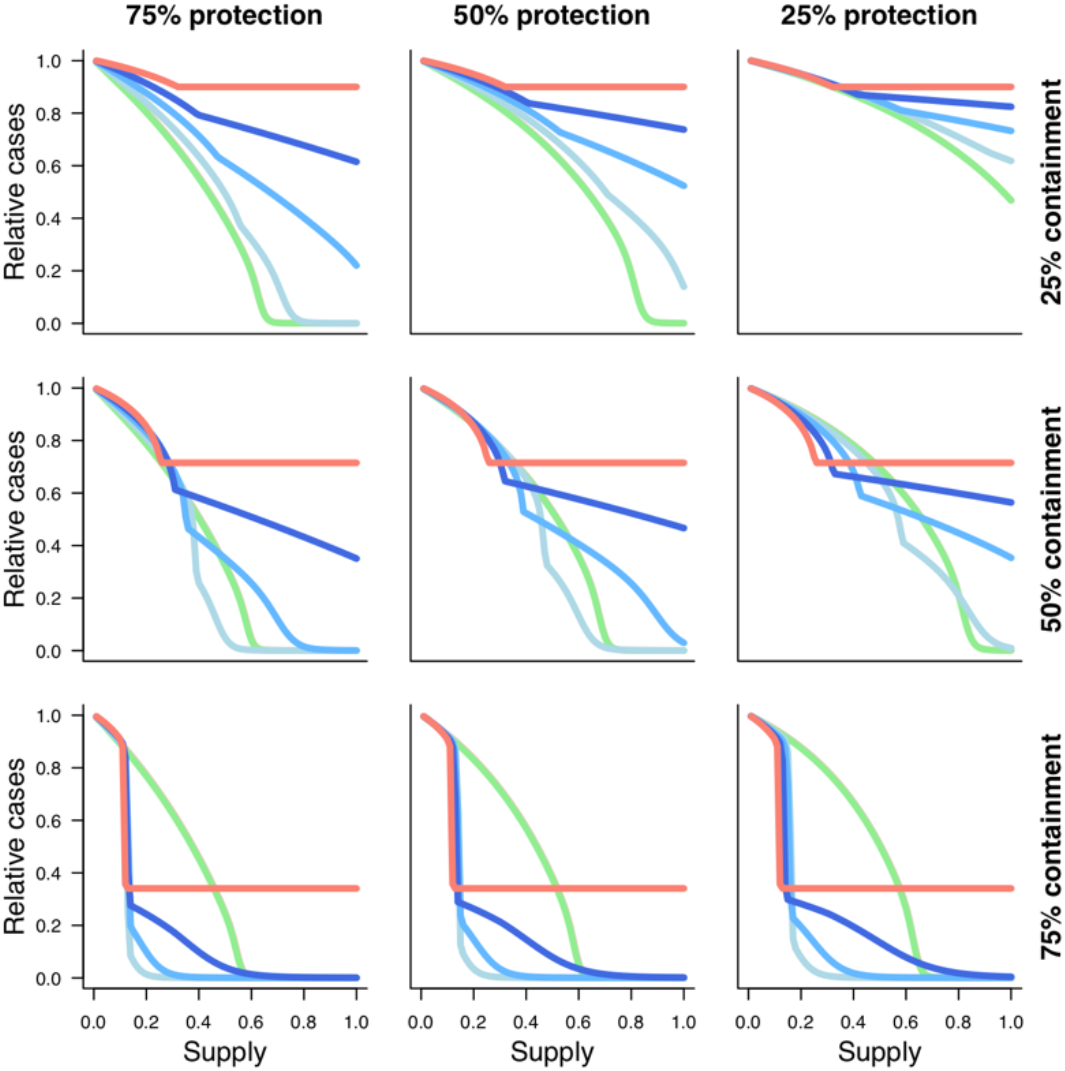
Reduction in total infections under each of the described resource allocation strategy for a range of resource availability levels. Corresponds to Figure 2. Each panel represents intervention effectiveness in terms of relative susceptibility and transmissibility, with the bottom left panel denoting the most effective intervention (75% reduction in susceptibility and transmissibility) and the top right panel representing the least effective intervention (25% reduction in susceptibility and transmissibility). Resources are provided naïvely (pink), prioritized to the elderly (green), saved for detected cases (red), or balanced at different levels between healthy individuals, prioritizing the elderly, and detected cases (blue). 30% of cases are assumed to be undetected. See *Methods* for further details.

**Figure S2.**
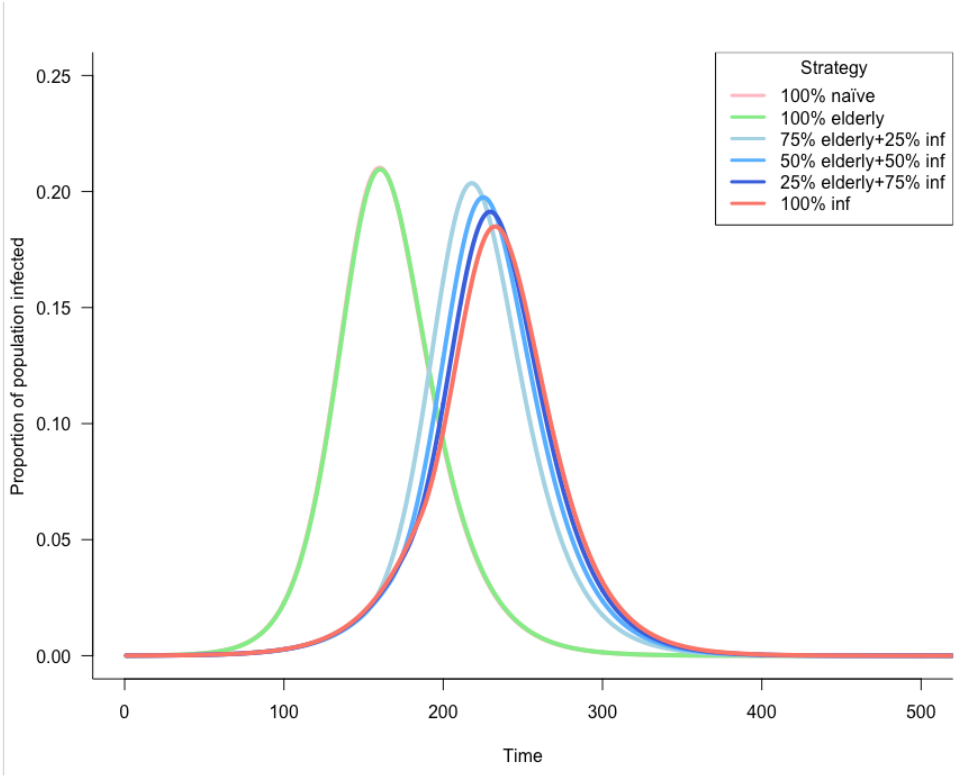
Optimizing mask distribution strategy can delay epidemic peak. We demonstrate that, even low coverage (5% of the population) of a mask offering no protection and 40% containment, the epidemic peak can be delayed when retaining resources for detected cases.

**Figure S3.**
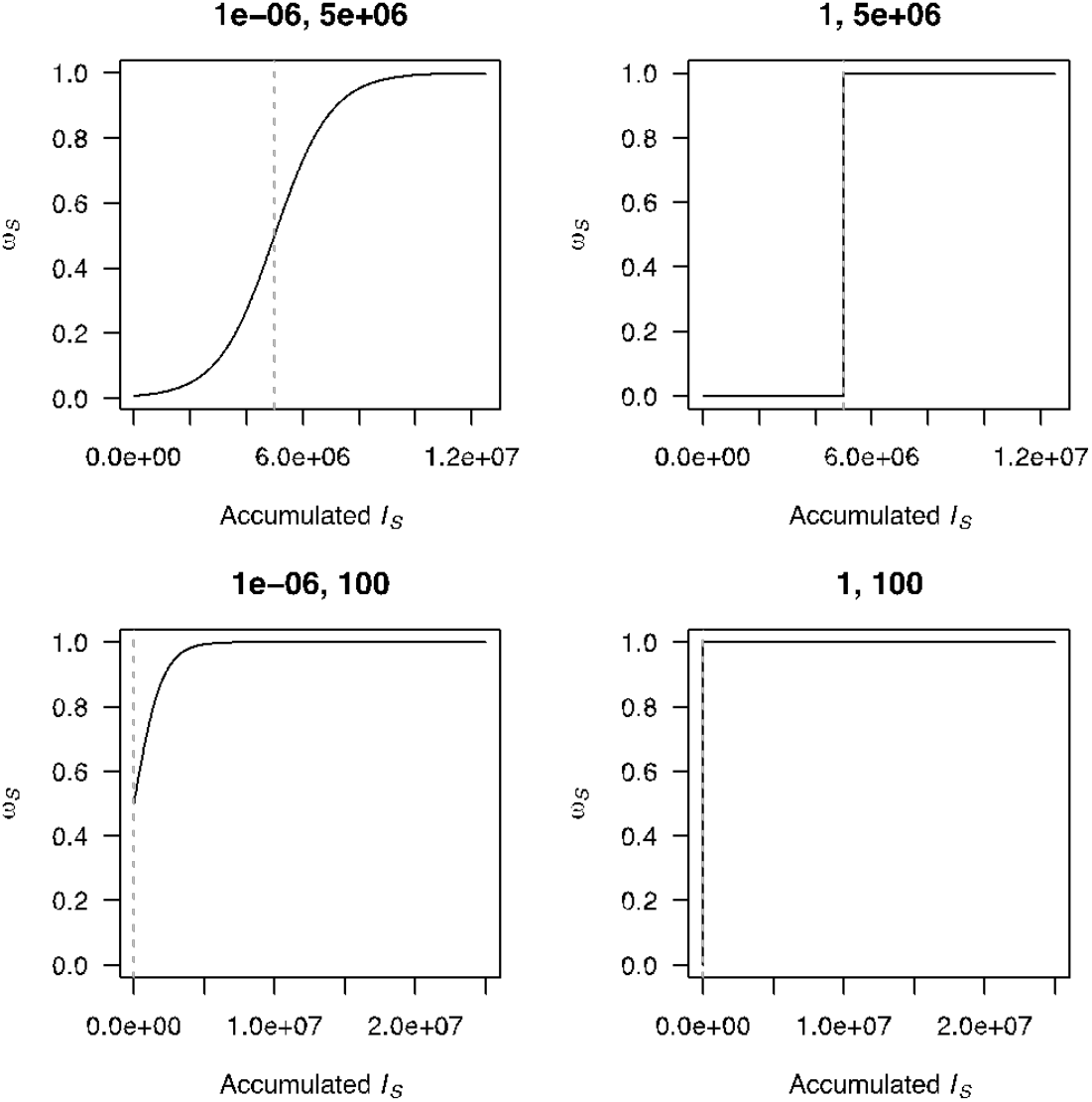
Different parameterizations of demand dynamics. Different values of *k*_*1*_ and *k*_*2*_, provided as the title for each panel, result in different demand responses to increasing numbers of reported cases. A panic buying scenario can be approximated in the bottom right panel, where demand is maximal very early in the epidemic. A gradual increase in demand is shown in the top left panel, in which 50% of the population seek masks if and when cases reach 5 million.

**Figure S4.**
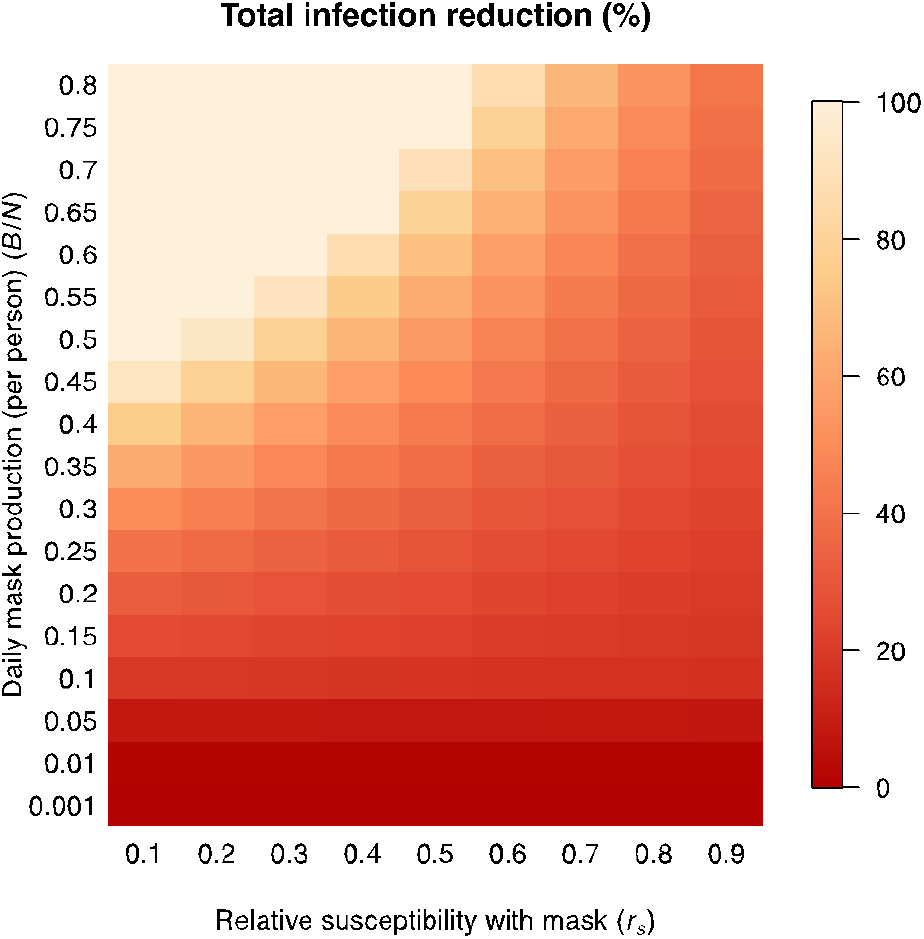
Impact of mask production rates on final number of infections. For different levels of mask protection and mask production, to reduction in total numbers of infections.

**Figure S5.**
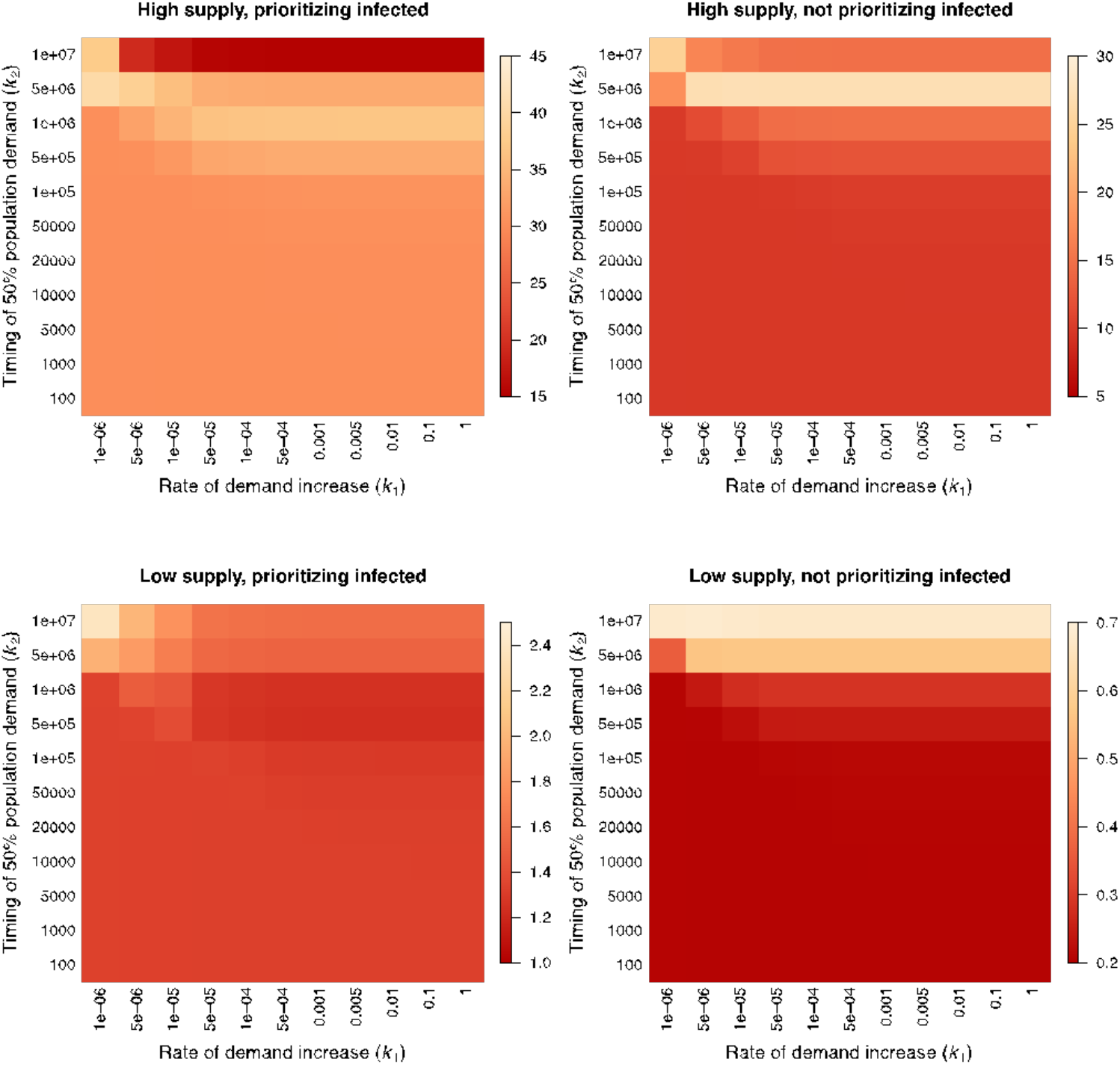
Levels of reduction in total infection numbers compared to no mask condition under different demand functions. The level of total infection reduction (%) varies with the timing of 50% population demand on masks (*k*_*1*_) and the rate of demand increase (*k*_*2*_) given high (top; *B*/*N*= 30%) or low bottom; *B*/*N*= 1%) mask production when prioritizing (left) or not prioritizing (right) masks for infectious cases. Generally, ‘panic buying’ is detrimental and prioritizing to infectious cases (setting *ω*_*S*_>*ω*_*A*_) is beneficial.

**Figure S6.**
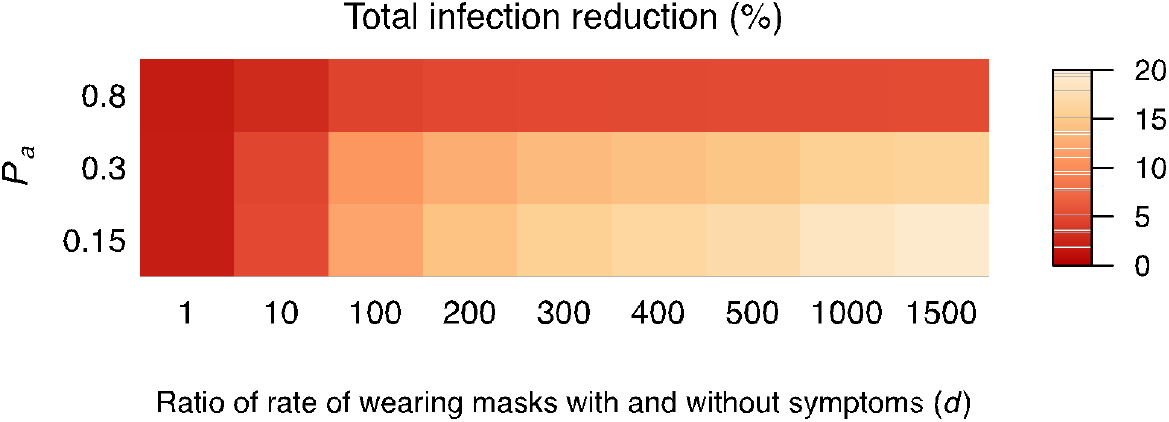
**The effect of prioritizing masks to infectious cases is less apparent if the proportion of asymptomatic infections is high.**

**Figure S7.**
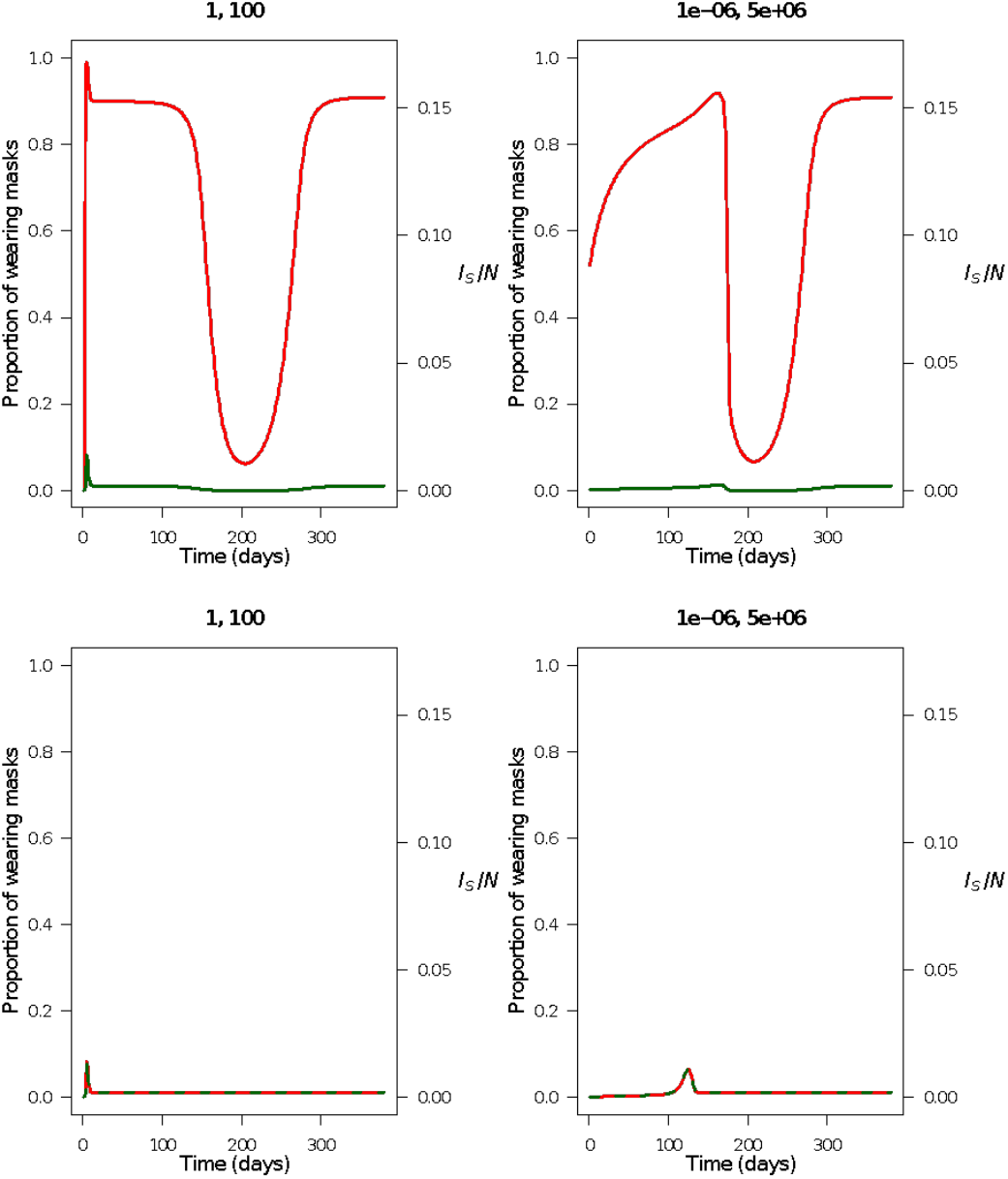
Low mask production rate can limit the advantage of building up supplies. Epidemic curves (pink) under a ‘panic buying’ demand curve (left), and a more gradual managed demand curve (right) when prioritizing (top) and not prioritizing (bottom) masks for infectious cases. Demand is shown as a dashed grey line, while the relative available mask supply is shown in blue. The proportion of the susceptible and symptomatically infected population wearing masks are shown as green and red lines, respectively. (*k*_*1*_, *k*_*2*_) are shown on the top of each subfigure. While supplies are built up in the early phase of the epidemic, shortage still occurs during the outbreak if mask production rate is low (*B*/*N* = 1% here).

## SUPPLEMENTARY TABLE

| Parameter | Definition | Baseline value | References |
| --- | --- | --- | --- |
| $R_0$ | Basic reproduction number for symptomatic individuals | 2.5 | [6]<br>[34]<br>[26] |
| $1/\alpha_1$ | Time from exposed to pre-symptomatic | 1 | [6] |
| $1/\alpha_2$ | Time from pre-symptomatic to symptomatic/ asymptomatic | 5 | [6] |
| $P_a$ | The proportion of asymptomatic infections | 30% (varied in simulations) | [30] |
| $1/\gamma_A$ | Time to recovery for asymptomatic infections | 14 | [6][27] |
| $1/\gamma_S$ | Time to recovery for symptomatic infections | 14 | [6][27] |
| $r_t$ | Relative transmissibility with mask | Varied in simulations<br>(=0.5 if not specified) | |
| $r_s$ | Relative susceptibility with mask | Varied in simulations<br>(=0.6 if not specified) | |
| **Resource allocation model** |  |  |  |
| $N_{70}$ | Proportion of people age 70+ | 7.6% | Taiwan as an example:<br>census.dgb<br>as.gov.tw |
|  | Death rate among symptomatics in 70+<br>Death rate among symptomatics in <70 | 9.7%<br>1.3% | [32] |
|  | Detection rate of non-severe infections | 44% [Japan] | [35] |
| ***Supply & demand model*** |  |  |  |
| $1/\mu$ | Average time of wearing disposable mask | 1 | |
| $B$ | Daily mask production | Varied in simulations | |
| $d$ | The ratio of rate of wearing mask between symptomatic infected individuals and others ($\omega_S/\omega_A$) | Varied in simulations<br>(=1000 if not specified) | |

## Notes

### Competing Interest Statement

The authors have declared no competing interest.

